# Anesthesiologists, Certified Registered Nurse Anesthetists and Anesthesiology Assistants: Overvaluation By Providers Or Undervaluation By Payers

**DOI:** 10.1101/2023.03.26.23287753

**Authors:** Deepak Gupta

## Abstract

**Background:** Considering that National Health Expenditures (NHE) in the United States (U.S.) have mounted to 4,124 billion dollars with 593.1 billion dollars (14.4%) spent on physician services in 2020, it will be interesting to see how specialties fared in terms of percentage of billed dollars approved to reflect on how NHE would have been faring if 100% billed dollars were approved.

**Objectives:** Whether (and by how much), as compared to other healthcare providers in the U.S., anesthesia providers (anesthesiologists, certified registered nurse anesthetists (CRNAs) and anesthesiology assistants) are being differently reimbursed in terms of approved dollars by Centers for Medicare & Medicaid Services (CMS) as percentages of dollars billed by providers. Materials and Methods: The source of data about approved dollars by CMS as percentages of dollars billed by providers is Denniston Data Inc. at Provider Ranking System (PRS) which provided deidentified cumulative tabulated data sourced from CMS public access reimbursement data for the years 2012-2020 to the author who, as a private citizen, had enrolled for annually renewed subscription to PRS.

**Results:** Among the 73 specialties whose data was complete for all the nine years (2012-2020), the top three specialties with highest percentages of billed dollars which were approved (56% to 76%) were almost consistently chiropractic, optometry and clinical psychologist respectively and anesthesiology assistants, CRNAs and anesthesiologists respectively were consistently the bottom three specialties with lowest percentages of billed dollars which were approved (11% to 18%).

**Conclusion:** During 2012-2020, anesthesia providers got 11%-18% of their billed dollars approved by CMS and yet anesthesiologists averaging almost top 1% wages (99th percentile) nationally with CRNAs averaging top 5% wages (95th percentile) nationally pose the question what their wages would have been if their billed dollars were 100% approved by CMS.

## Introduction

As an anesthesiologist who have had the opportunity to train and work in two different worlds [1], the author has always wondered about the differences between developed world and developing world in terms of anesthesia practice. There may already be enormous clinical practice literature documenting their differences due to their different resources. However, there may still be limited healthcare economics literature documenting their differences due to their different reimbursements. This comparison may still be very difficult to decipher because wage data for practitioners may not be officially available for India. However, estimated wage data by occupation from United States (U.S.) Bureau of Labor Statistics (BLS) is formally published as Occupational Employment and Wage Statistics (OEWS) annually [2-11]. Although data based on tax returns officially filed as Form 1040 (U.S. Individual Income Tax Return) annually with the Internal Revenue Service (IRS) may be the best approximated wage data by occupation, publicly available BLS OEWS may be the next best thing in the absence of publicly available deidentified IRS Form 1040 data accumulated for entire population and assorted per occupation. Considering that U.S. National Health Expenditures (NHE) have mounted to 4,124 billion dollars representing 19.7% of U.S. Gross Domestic Product (GDP) with 593.1 billion dollars (14.4%) spent on physician services in 2020 [12], it will be interesting to see how specialties fared in terms of percentage of billed dollars approved just like Cost-to-Charge Ratios [13-15], locally and nationally, depicting how hospital-related NHE (1,270.1 billion dollars (30.8%) spent on hospital care in 2020) and thus GDP would have been faring if 100% billed dollars were approved.

To compare available BLS OEWS with public access reimbursement data from the largest healthcare payer in the U.S., Centers for Medicare & Medicaid Services (CMS) as tabulated by Denniston Data Inc. at Provider Ranking System (PRS) [16-17], the objective for this project is to present whether (and by how much), as compared to other healthcare providers in the U.S., anesthesia providers (anesthesiologists, certified registered nurse anesthetists (CRNAs) and anesthesiology assistants) are being differently reimbursed in terms of approved dollars by CMS as percentages of dollars billed by providers.

## Materials and Methods

The data is hereby being presented after institutional review board (IRB) determined that this project did not meet the definition of Human Participant Research subject to IRB oversight and review because of public use datasets with de-Identified private information ensuring the project neither meeting the regulatory definition of research per Code of Federal Regulations (CFR) 45 CFR 46.102(l) nor meeting the regulatory definition of human subjects/participants per 45 CFR 46.102(e) [18]. The source of data about approved dollars by CMS as percentages of dollars billed by providers is Denniston Data Inc. at PRS which provided deidentified cumulative tabulated data sourced from CMS public access reimbursement data for the years 2012-2020 (See Supplementary Excel File) to the author who, as a private citizen, had enrolled for annually renewed subscription to PRS. For the years 2018-2020, annual data for billed dollars, approved dollars, codes billed, services billed, patients cared and total providers were available whereafter the data was converted into tables of billed dollars per provider, approved dollars per provider, codes billed per provider, services billed per provider and patients cared per provider. For the years 2012-2017, annual data for billed dollars, approved dollars, codes billed, services billed, and patients cared were available but in the absence of annual data about total providers for the years 2012-2017, the tabulated data for the whole data period 2012-2020 was limited to table of absolute billed dollars, table of absolute approved dollars, and table of percentages of billed dollars which were approved. Publicly available BLS OEWS data used to compare was May 2021 estimates about employment and annual mean wage averaged over three-year period and rolling six-panels (May 2021, November 2020, May 2020, November 2019, May 2019 and November 2018 [4]) which were compared to 2018-2020 period PRS data averaged annually with it being the closest three-year period of PRS data wherein approved dollars’ data and total providers’ data were available.

## Results

Among a total of 102 specialties’ plus non-specialties’ cumulative data provided by PRS (See Supplementary Excel File), 73 specialties’ data was complete for all the nine years (2012-2020) which were thereafter tabulated as Tables 1-3 as well as charted as Figures 1-3 (also see interactive charts in Supplementary Excel File) for absolute billed dollars, absolute approved dollars and percentages of billed dollars which were approved, respectively. Herein, it can be seen that the top three specialties with highest absolute billed dollars were almost consistently internal medicine, diagnostic radiology and hematology-oncology respectively while the top three specialties with highest absolute approved dollars were almost consistently internal medicine, ophthalmology and hematology-oncology respectively. However, the top three specialties with highest percentages of billed dollars which were approved were almost consistently chiropractic, optometry and clinical psychologist respectively. Regarding anesthesia providers, anesthesiology assistants’ absolute billed dollars, absolute approved dollars and percentages of billed dollars which were approved hovered around the bottom while anesthesiologists from being near the top in absolute billed dollars and CRNAs from being near the middle in absolute billed dollars gradually bottomed out when the data moved to absolute approved dollars and thence to percentages of billed dollars which were approved thus consistently making the bottom three specialties in terms of percentages of billed dollars which were approved (presented as broad colored lines in Figures 1-3 with grey accent 3 color, olive orange yellow color and gold accent 4 color representing anesthesiologists, CRNAs and anesthesiology assistants respectively). For 2018-2020 period, Tables 4-6 respectively documented among each of the 73 specialties how providers on an average annually billed CMS for dollars, got annually approved dollars from CMS for codes and services annually billed for patients cared annually and where the anesthesia providers (anesthesiologists, CRNAs and anesthesiology assistants) stood in comparison to other healthcare providers. Table 7 depicts the comparison between PRS data and BLS data only for the 23 specialties concurrently available on both databases wherein also such drawn comparisons could only be metaphorical (either subjective or qualitative as neither quantitative nor objective) due to discrepancies between inclusion and exclusion methods among PRS data and BLS data.

**Table 1:** Absolute Billed Dollars (In Millions) Per Specialty For 2012-2020 Period.

| Specialty Name | YEAR 2012 | YEAR 2013 | YEAR 2014 | YEAR 2015 | YEAR 2016 | YEAR 2017 | YEAR 2018 | YEAR 2019 | YEAR 2020 |
| --- | --- | --- | --- | --- | --- | --- | --- | --- | --- |
| Addiction Medicine | 11 | 14 | 15 | 17 | 20 | 21 | 25 | 25 | 20 |
| Allergy/ Immunology | 447 | 478 | 515 | 593 | 652 | 692 | 761 | 868 | 888 |
| Anesthesiology | 11,168 | 12,060 | 12,675 | 13,308 | 14,332 | 14,998 | 15,661 | 16,217 | 14,011 |
| Anesthesiology Assistant | 137 | 169 | 211 | 250 | 287 | 325 | 372 | 403 | 364 |
| Audiologist | 154 | 168 | 172 | 178 | 191 | 199 | 205 | 215 | 177 |
| Cardiac Surgery | 1,307 | 1,261 | 1,245 | 1,166 | 1,147 | 1,117 | 1,093 | 1,057 | 845 |
| Cardiology | 18,271 | 17,833 | 17,819 | 16,081 | 16,062 | 15,966 | 15,585 | 15,719 | 13,531 |
| Certified Clinical Nurse Specialist | 120 | 135 | 139 | 143 | 154 | 161 | 161 | 157 | 140 |
| Certified Nurse Midwife | 5 | 6 | 6 | 6 | 8 | 8 | 9 | 9 | 7 |
| Certified Registered Nurse Anesthetist | 5,354 | 5,940 | 6,435 | 7,146 | 7,953 | 8,499 | 9,199 | 9,863 | 8,696 |
| Chiropractic | 977 | 975 | 991 | 1,006 | 1,014 | 998 | 1,019 | 1,044 | 880 |
| Clinical Cardiac Electrophysiology | 948 | 1,059 | 1,203 | 1,386 | 1,612 | 1,789 | 1,956 | 2,265 | 2,158 |
| Colorectal Surgery (Proctology) | 444 | 463 | 490 | 508 | 538 | 563 | 559 | 565 | 477 |
| Critical Care (Intensivists) | 708 | 725 | 772 | 853 | 941 | 1,023 | 1,068 | 1,162 | 1,175 |
| Dermatology | 5,633 | 5,969 | 6,269 | 6,612 | 7,089 | 7,349 | 7,685 | 8,241 | 7,465 |
| Diagnostic Radiology | 18,328 | 18,418 | 18,570 | 19,553 | 20,493 | 21,021 | 21,918 | 23,268 | 21,257 |
| Emergency Medicine | 11,847 | 12,819 | 14,001 | 15,355 | 16,592 | 17,334 | 17,795 | 18,427 | 15,435 |
| Endocrinology | 996 | 1,049 | 1,078 | 1,143 | 1,218 | 1,277 | 1,354 | 1,424 | 1,310 |
| Family Practice | 11,772 | 12,153 | 12,514 | 13,349 | 13,932 | 14,423 | 14,432 | 14,772 | 13,117 |
| Gastroenterology | 5,966 | 6,131 | 6,106 | 6,252 | 6,632 | 6,800 | 7,020 | 7,307 | 6,253 |
| General Practice | 1,024 | 986 | 925 | 860 | 864 | 836 | 773 | 741 | 648 |
| General Surgery | 6,586 | 6,686 | 6,647 | 6,703 | 6,849 | 6,745 | 6,787 | 6,819 | 5,927 |
| Geriatric Medicine | 422 | 424 | 414 | 433 | 421 | 410 | 417 | 432 | 382 |
| Geriatric Psychiatry | 20 | 25 | 27 | 32 | 36 | 40 | 39 | 37 | 33 |
| Gynecological Oncology | 446 | 469 | 457 | 467 | 472 | 487 | 486 | 517 | 521 |
| Hand Surgery | 543 | 562 | 599 | 656 | 730 | 791 | 860 | 933 | 822 |
| Hematology | 639 | 643 | 631 | 569 | 615 | 619 | 711 | 757 | 716 |
| Hematology-Oncology | 14,054 | 13,890 | 14,030 | 14,886 | 16,311 | 17,263 | 18,864 | 20,347 | 20,708 |
| Hospice and Palliative Care | 33 | 49 | 62 | 82 | 98 | 115 | 133 | 152 | 157 |
| Infectious Disease | 1,413 | 1,468 | 1,527 | 1,613 | 1,660 | 1,712 | 1,784 | 1,838 | 1,871 |
| Internal Medicine | 23,197 | 23,630 | 24,053 | 25,100 | 25,903 | 25,970 | 24,575 | 24,903 | 22,463 |
| Interventional Pain Management | 1,820 | 1,988 | 1,933 | 1,834 | 1,866 | 1,964 | 2,043 | 2,036 | 1,810 |
| Interventional Radiology | 1,012 | 1,254 | 1,348 | 1,438 | 1,622 | 1,712 | 1,929 | 2,191 | 2,173 |
| Licensed Clinical Social Worker | 497 | 552 | 596 | 648 | 727 | 792 | 844 | 899 | 829 |
| Maxillofacial Surgery | 45 | 47 | 46 | 47 | 52 | 55 | 55 | 57 | 50 |
| Medical Oncology | 4,288 | 4,321 | 4,357 | 4,554 | 5,032 | 5,407 | 6,010 | 6,678 | 6,813 |
| Nephrology | 4,986 | 5,141 | 5,158 | 5,255 | 5,439 | 5,145 | 5,150 | 5,219 | 5,018 |
| Neurology | 3,920 | 3,912 | 4,105 | 4,370 | 4,657 | 4,926 | 5,338 | 5,663 | 5,043 |
| Neuropsychiatry | 21 | 21 | 26 | 28 | 28 | 32 | 27 | 29 | 21 |
| Neurosurgery | 3,229 | 3,348 | 3,479 | 3,643 | 3,829 | 3,866 | 3,918 | 3,923 | 3,535 |
| Nuclear Medicine | 302 | 302 | 306 | 316 | 345 | 365 | 389 | 409 | 416 |
| Nurse Practitioner | 4,005 | 4,822 | 5,718 | 6,998 | 8,406 | 10,058 | 11,790 | 14,001 | 14,431 |
| Obstetrics & Gynecology | 1,526 | 1,528 | 1,507 | 1,499 | 1,541 | 1,531 | 1,549 | 1,575 | 1,320 |
| Occupational Therapist in Private Practice | 301 | 298 | 334 | 378 | 423 | 490 | 579 | 690 | 632 |
| Ophthalmology | 15,827 | 17,088 | 17,531 | 18,401 | 19,447 | 19,853 | 20,662 | 21,825 | 19,385 |
| Optometry | 1,348 | 1,459 | 1,539 | 1,643 | 1,772 | 1,874 | 1,966 | 2,083 | 1,702 |
| Oral Surgery (Dentist only) | 55 | 57 | 63 | 73 | 93 | 127 | 133 | 127 | 113 |
| Orthopedic Surgery | 12,917 | 13,445 | 13,705 | 14,046 | 14,696 | 14,881 | 15,248 | 15,515 | 13,356 |
| Osteopathic Manipulative Medicine | 107 | 108 | 115 | 120 | 127 | 132 | 136 | 144 | 128 |
| Otolaryngology | 2,663 | 2,906 | 3,073 | 3,219 | 3,342 | 3,423 | 3,460 | 3,577 | 2,906 |
| Pain Management | 821 | 1,047 | 1,280 | 1,636 | 1,841 | 1,990 | 2,146 | 2,367 | 2,129 |
| Pathology | 4,125 | 4,168 | 4,082 | 4,189 | 4,354 | 4,504 | 4,762 | 4,960 | 4,416 |
| Pediatric Medicine | 186 | 177 | 196 | 200 | 208 | 206 | 196 | 214 | 188 |
| Peripheral Vascular Disease | 66 | 66 | 60 | 54 | 53 | 65 | 67 | 69 | 60 |
| Physical Medicine and Rehabilitation | 2,628 | 2,682 | 2,873 | 3,066 | 3,238 | 3,408 | 3,602 | 3,774 | 3,341 |
| Physical Therapist in Private Practice | 4,165 | 4,330 | 4,855 | 5,458 | 6,145 | 6,762 | 7,454 | 8,415 | 6,922 |
| Physician Assistant | 4,761 | 5,461 | 6,116 | 6,963 | 8,054 | 9,015 | 10,002 | 11,031 | 10,226 |
| Plastic and Reconstructive Surgery | 1,190 | 1,230 | 1,275 | 1,317 | 1,389 | 1,434 | 1,490 | 1,549 | 1,405 |
| Podiatry | 3,312 | 3,391 | 3,463 | 3,544 | 3,721 | 3,813 | 4,004 | 4,098 | 3,756 |
| Preventive Medicine | 35 | 52 | 59 | 53 | 53 | 56 | 51 | 47 | 40 |
| Psychiatry | 2,122 | 2,279 | 2,285 | 2,245 | 2,265 | 2,247 | 2,213 | 2,197 | 1,972 |
| Psychologist, Clinical | 23 | 22 | 18 | 20 | 1,133 | 1,160 | 1,207 | 1,223 | 1,097 |
| Pulmonary Disease | 3,705 | 3,805 | 3,880 | 4,011 | 4,107 | 4,165 | 4,163 | 4,168 | 3,775 |
| Radiation Oncology | 7,027 | 7,047 | 7,168 | 7,150 | 7,199 | 7,297 | 7,528 | 7,810 | 7,223 |
| Registered Dietitian or Nutrition Profess | 24 | 26 | 28 | 32 | 34 | 35 | 37 | 41 | 35 |
| Rheumatology | 2,945 | 3,167 | 3,432 | 3,817 | 4,326 | 4,779 | 5,389 | 6,154 | 6,462 |
| Sleep Medicine | 0 | 27 | 48 | 73 | 96 | 134 | 145 | 162 | 141 |
| Speech Language Pathologist | 30 | 23 | 28 | 42 | 51 | 67 | 84 | 104 | 91 |
| Sports Medicine | 84 | 133 | 186 | 251 | 314 | 382 | 422 | 481 | 451 |
| Surgical Oncology | 239 | 251 | 274 | 289 | 319 | 326 | 344 | 373 | 337 |
| Thoracic Surgery | 1,251 | 1,280 | 1,272 | 1,308 | 1,318 | 1,335 | 1,333 | 1,352 | 1,200 |
| Urology | 6,341 | 6,393 | 6,312 | 6,342 | 6,516 | 6,604 | 6,728 | 6,986 | 6,258 |
| Vascular Surgery | 3,054 | 3,331 | 3,466 | 3,584 | 3,865 | 3,872 | 4,064 | 4,384 | 3,980 |
| TOTAL | 249,972 | 259,644 | 268,199 | 280,462 | 298,871 | 309,838 | 321,963 | 339,081 | 307,643 |

Source of data: CMS public access data tabulated by Denniston Data Inc. at PRS [16-17]

**Table 2:** Absolute Approved Dollars (In Millions) Per Specialty For 2012-2020 Period.

| Specialty Name | YEAR 2012 | YEAR 2013 | YEAR 2014 | YEAR 2015 | YEAR 2016 | YEAR 2017 | YEAR 2018 | YEAR 2019 | YEAR 2020 |
| --- | --- | --- | --- | --- | --- | --- | --- | --- | --- |
| Addiction Medicine | 6 | 8 | 8 | 8 | 9 | 10 | 12 | 12 | 9 |
| Allergy/ Immunology | 262 | 280 | 292 | 326 | 356 | 377 | 396 | 429 | 431 |
| Anesthesiology | 2,003 | 2,080 | 2,111 | 2,131 | 2,134 | 2,142 | 2,129 | 2,090 | 1,796 |
| Anesthesiology Assistant | 22 | 25 | 29 | 32 | 34 | 37 | 41 | 43 | 38 |
| Audiologist | 59 | 60 | 61 | 62 | 66 | 69 | 71 | 73 | 58 |
| Cardiac Surgery | 367 | 349 | 346 | 324 | 311 | 300 | 289 | 273 | 211 |
| Cardiology | 6,880 | 6,603 | 6,640 | 5,988 | 5,866 | 5,772 | 5,606 | 5,559 | 4,781 |
| Certified Clinical Nurse Specialist | 57 | 65 | 67 | 67 | 69 | 70 | 70 | 66 | 58 |
| Certified Nurse Midwife | 3 | 3 | 3 | 3 | 4 | 4 | 4 | 4 | 3 |
| Certified Registered Nurse Anesthetist | 978 | 1,043 | 1,100 | 1,133 | 1,160 | 1,195 | 1,219 | 1,241 | 1,066 |
| Chiropractic | 689 | 684 | 750 | 745 | 735 | 711 | 724 | 738 | 622 |
| Clinical Cardiac Electrophysiology | 301 | 354 | 416 | 482 | 558 | 609 | 648 | 708 | 674 |
| Colorectal Surgery (Proctology) | 158 | 162 | 163 | 165 | 167 | 167 | 165 | 166 | 143 |
| Critical Care (Intensivists) | 286 | 285 | 299 | 321 | 339 | 357 | 367 | 379 | 378 |
| Dermatology | 3,183 | 3,284 | 3,345 | 3,385 | 3,518 | 3,577 | 3,665 | 3,803 | 3,391 |
| Diagnostic Radiology | 5,032 | 4,836 | 4,777 | 4,870 | 5,056 | 5,233 | 5,375 | 5,502 | 4,885 |
| Emergency Medicine | 3,001 | 3,012 | 3,135 | 3,216 | 3,227 | 3,220 | 3,140 | 3,067 | 2,568 |
| Endocrinology | 529 | 549 | 548 | 563 | 583 | 604 | 624 | 629 | 576 |
| Family Practice | 6,455 | 6,548 | 6,557 | 6,850 | 6,831 | 6,836 | 6,720 | 6,572 | 5,804 |
| Gastroenterology | 1,951 | 1,959 | 1,901 | 1,909 | 1,893 | 1,876 | 1,907 | 1,928 | 1,681 |
| General Practice | 550 | 517 | 478 | 444 | 432 | 413 | 382 | 358 | 308 |
| General Surgery | 2,175 | 2,169 | 2,128 | 2,098 | 2,073 | 2,012 | 1,993 | 1,966 | 1,699 |
| Geriatric Medicine | 235 | 233 | 224 | 227 | 215 | 205 | 201 | 198 | 175 |
| Geriatric Psychiatry | 12 | 14 | 16 | 18 | 19 | 20 | 20 | 19 | 16 |
| Gynecological Oncology | 134 | 142 | 140 | 142 | 144 | 149 | 148 | 155 | 154 |
| Hand Surgery | 158 | 171 | 179 | 193 | 211 | 226 | 242 | 260 | 229 |
| Hematology | 277 | 265 | 249 | 220 | 235 | 239 | 271 | 269 | 257 |
| Hematology-Oncology | 6,044 | 5,775 | 5,724 | 5,936 | 6,551 | 6,967 | 7,562 | 7,824 | 7,871 |
| Hospice and Palliative Care | 15 | 21 | 27 | 36 | 43 | 49 | 56 | 61 | 64 |
| Infectious Disease | 710 | 721 | 748 | 769 | 770 | 780 | 794 | 791 | 803 |
| Internal Medicine | 12,115 | 12,106 | 12,140 | 12,400 | 12,229 | 11,691 | 11,008 | 10,640 | 9,452 |
| Interventional Pain Management | 522 | 569 | 523 | 503 | 482 | 497 | 502 | 493 | 441 |
| Interventional Radiology | 237 | 293 | 309 | 323 | 368 | 395 | 445 | 508 | 502 |
| Licensed Clinical Social Worker | 248 | 282 | 330 | 353 | 394 | 420 | 456 | 477 | 438 |
| Maxillofacial Surgery | 18 | 18 | 17 | 16 | 17 | 17 | 17 | 17 | 15 |
| Medical Oncology | 1,768 | 1,716 | 1,697 | 1,758 | 1,954 | 2,084 | 2,311 | 2,469 | 2,470 |
| Nephrology | 2,241 | 2,265 | 2,293 | 2,307 | 2,349 | 2,292 | 2,290 | 2,267 | 2,164 |
| Neurology | 1,829 | 1,767 | 1,802 | 1,857 | 1,906 | 1,955 | 2,033 | 2,058 | 1,808 |
| Neuropsychiatry | 10 | 11 | 13 | 12 | 12 | 13 | 10 | 10 | 9 |
| Neurosurgery | 713 | 730 | 759 | 784 | 807 | 798 | 804 | 804 | 711 |
| Nuclear Medicine | 87 | 86 | 95 | 99 | 109 | 120 | 128 | 130 | 131 |
| Nurse Practitioner | 1,673 | 1,972 | 2,314 | 2,760 | 3,207 | 3,751 | 4,322 | 4,950 | 5,170 |
| Obstetrics & Gynecology | 631 | 617 | 608 | 591 | 594 | 578 | 576 | 572 | 484 |
| Occupational Therapist in Private Practice | 180 | 162 | 172 | 192 | 208 | 233 | 269 | 310 | 290 |
| Ophthalmology | 7,758 | 8,223 | 8,413 | 8,645 | 9,003 | 9,135 | 9,503 | 9,778 | 8,625 |
| Optometry | 1,026 | 1,088 | 1,128 | 1,163 | 1,222 | 1,252 | 1,297 | 1,342 | 1,075 |
| Oral Surgery (Dentist only) | 26 | 26 | 27 | 31 | 39 | 54 | 59 | 58 | 51 |
| Orthopedic Surgery | 3,893 | 3,965 | 3,877 | 3,910 | 3,999 | 3,983 | 4,009 | 4,097 | 3,543 |
| Osteopathic Manipulative Medicine | 53 | 50 | 50 | 51 | 53 | 53 | 54 | 56 | 52 |
| Otolaryngology | 1,118 | 1,202 | 1,208 | 1,224 | 1,237 | 1,238 | 1,249 | 1,256 | 1,027 |
| Pain Management | 243 | 298 | 344 | 427 | 450 | 486 | 516 | 552 | 516 |
| Pathology | 1,296 | 1,214 | 1,130 | 1,192 | 1,257 | 1,289 | 1,360 | 1,370 | 1,227 |
| Pediatric Medicine | 81 | 73 | 82 | 79 | 81 | 79 | 78 | 81 | 69 |
| Peripheral Vascular Disease | 22 | 21 | 16 | 14 | 15 | 20 | 23 | 24 | 20 |
| Physical Medicine and Rehabilitation | 1,146 | 1,126 | 1,146 | 1,194 | 1,211 | 1,222 | 1,255 | 1,269 | 1,121 |
| Physical Therapist in Private Practice | 2,362 | 2,263 | 2,425 | 2,669 | 2,923 | 3,152 | 3,403 | 3,717 | 3,046 |
| Physician Assistant | 1,218 | 1,385 | 1,547 | 1,761 | 1,996 | 2,245 | 2,471 | 2,716 | 2,559 |
| Plastic and Reconstructive Surgery | 371 | 373 | 376 | 374 | 379 | 377 | 385 | 384 | 344 |
| Podiatry | 2,020 | 2,061 | 2,046 | 2,028 | 2,041 | 2,047 | 2,129 | 2,156 | 1,987 |
| Preventive Medicine | 15 | 20 | 22 | 20 | 19 | 19 | 18 | 17 | 15 |
| Psychiatry | 1,151 | 1,258 | 1,290 | 1,235 | 1,198 | 1,153 | 1,120 | 1,081 | 961 |
| Psychologist, Clinical | 13 | 13 | 12 | 13 | 664 | 672 | 683 | 680 | 618 |
| Pulmonary Disease | 1,822 | 1,840 | 1,837 | 1,846 | 1,817 | 1,805 | 1,777 | 1,730 | 1,546 |
| Radiation Oncology | 2,017 | 1,908 | 1,994 | 1,966 | 1,906 | 1,915 | 1,966 | 1,997 | 1,862 |
| Registered Dietitian or Nutrition Profess | 13 | 14 | 15 | 16 | 16 | 16 | 17 | 19 | 16 |
| Rheumatology | 1,605 | 1,733 | 1,858 | 2,087 | 2,370 | 2,620 | 2,908 | 3,060 | 3,150 |
| Sleep Medicine | 0 | 10 | 18 | 27 | 36 | 48 | 54 | 57 | 51 |
| Speech Language Pathologist | 19 | 14 | 17 | 25 | 29 | 35 | 44 | 55 | 48 |
| Sports Medicine | 27 | 43 | 58 | 81 | 99 | 120 | 133 | 157 | 143 |
| Surgical Oncology | 70 | 72 | 78 | 80 | 87 | 89 | 94 | 96 | 87 |
| Thoracic Surgery | 339 | 336 | 348 | 357 | 356 | 359 | 358 | 354 | 313 |
| Urology | 2,344 | 2,310 | 2,253 | 2,264 | 2,308 | 2,271 | 2,266 | 2,272 | 2,038 |
| Vascular Surgery | 901 | 964 | 1,012 | 1,049 | 1,110 | 1,147 | 1,217 | 1,295 | 1,167 |
| TOTAL | 97,771 | 98,710 | 100,163 | 102,447 | 106,167 | 107,969 | 110,458 | 112,614 | 102,110 |

Source of data: CMS public access data tabulated by Denniston Data Inc. at PRS [16-17]

**Table 3:** Percentages Of Billed Dollars Which Were Approved For Each Specialty During 2012-2020 Period.

| Specialty Name | YEAR 2012 | YEAR 2013 | YEAR 2014 | YEAR 2015 | YEAR 2016 | YEAR 2017 | YEAR 2018 | YEAR 2019 | YEAR 2020 |
| --- | --- | --- | --- | --- | --- | --- | --- | --- | --- |
| Addiction Medicine | 55 | 52 | 51 | 49 | 46 | 47 | 48 | 46 | 46 |
| Allergy/ Immunology | 59 | 58 | 57 | 55 | 55 | 54 | 52 | 49 | 49 |
| Anesthesiology | 18 | 17 | 17 | 16 | 15 | 14 | 14 | 13 | 13 |
| Anesthesiology Assistant | 16 | 15 | 14 | 13 | 12 | 11 | 11 | 11 | 11 |
| Audiologist | 38 | 36 | 36 | 35 | 35 | 35 | 35 | 34 | 33 |
| Cardiac Surgery | 28 | 28 | 28 | 28 | 27 | 27 | 26 | 26 | 25 |
| Cardiology | 38 | 37 | 37 | 37 | 37 | 36 | 36 | 35 | 35 |
| Certified Clinical Nurse Specialist | 47 | 48 | 48 | 46 | 45 | 44 | 43 | 42 | 41 |
| Certified Nurse Midwife | 52 | 52 | 49 | 48 | 47 | 47 | 46 | 44 | 47 |
| Certified Registered Nurse Anesthetist | 18 | 18 | 17 | 16 | 15 | 14 | 13 | 13 | 12 |
| Chiropractic | 71 | 70 | 76 | 74 | 72 | 71 | 71 | 71 | 71 |
| Clinical Cardiac Electrophysiology | 32 | 33 | 35 | 35 | 35 | 34 | 33 | 31 | 31 |
| Colorectal Surgery (Proctology) | 36 | 35 | 33 | 33 | 31 | 30 | 30 | 29 | 30 |
| Critical Care (Intensivists) | 40 | 39 | 39 | 38 | 36 | 35 | 34 | 33 | 32 |
| Dermatology | 57 | 55 | 53 | 51 | 50 | 49 | 48 | 46 | 45 |
| Diagnostic Radiology | 27 | 26 | 26 | 25 | 25 | 25 | 25 | 24 | 23 |
| Emergency Medicine | 25 | 23 | 22 | 21 | 19 | 19 | 18 | 17 | 17 |
| Endocrinology | 53 | 52 | 51 | 49 | 48 | 47 | 46 | 44 | 44 |
| Family Practice | 55 | 54 | 52 | 51 | 49 | 47 | 47 | 44 | 44 |
| Gastroenterology | 33 | 32 | 31 | 31 | 29 | 28 | 27 | 26 | 27 |
| General Practice | 54 | 52 | 52 | 52 | 50 | 49 | 49 | 48 | 48 |
| General Surgery | 33 | 32 | 32 | 31 | 30 | 30 | 29 | 29 | 29 |
| Geriatric Medicine | 56 | 55 | 54 | 53 | 51 | 50 | 48 | 46 | 46 |
| Geriatric Psychiatry | 58 | 58 | 59 | 56 | 52 | 50 | 51 | 50 | 49 |
| Gynecological Oncology | 30 | 30 | 31 | 30 | 31 | 31 | 30 | 30 | 30 |
| Hand Surgery | 29 | 30 | 30 | 29 | 29 | 29 | 28 | 28 | 28 |
| Hematology | 43 | 41 | 39 | 39 | 38 | 39 | 38 | 35 | 36 |
| Hematology-Oncology | 43 | 42 | 41 | 40 | 40 | 40 | 40 | 38 | 38 |
| Hospice and Palliative Care | 45 | 44 | 44 | 44 | 43 | 43 | 42 | 40 | 41 |
| Infectious Disease | 50 | 49 | 49 | 48 | 46 | 46 | 45 | 43 | 43 |
| Internal Medicine | 52 | 51 | 50 | 49 | 47 | 45 | 45 | 43 | 42 |
| Interventional Pain Management | 29 | 29 | 27 | 27 | 26 | 25 | 25 | 24 | 24 |
| Interventional Radiology | 23 | 23 | 23 | 22 | 23 | 23 | 23 | 23 | 23 |
| Licensed Clinical Social Worker | 50 | 51 | 55 | 54 | 54 | 53 | 54 | 53 | 53 |
| Maxillofacial Surgery | 39 | 38 | 37 | 35 | 33 | 31 | 31 | 30 | 30 |
| Medical Oncology | 41 | 40 | 39 | 39 | 39 | 39 | 38 | 37 | 36 |
| Nephrology | 45 | 44 | 44 | 44 | 43 | 45 | 44 | 43 | 43 |
| Neurology | 47 | 45 | 44 | 43 | 41 | 40 | 38 | 36 | 36 |
| Neuropsychiatry | 49 | 50 | 52 | 44 | 41 | 39 | 37 | 35 | 42 |
| Neurosurgery | 22 | 22 | 22 | 22 | 21 | 21 | 21 | 20 | 20 |
| Nuclear Medicine | 29 | 28 | 31 | 31 | 32 | 33 | 33 | 32 | 31 |
| Nurse Practitioner | 42 | 41 | 40 | 39 | 38 | 37 | 37 | 35 | 36 |
| Obstetrics & Gynecology | 41 | 40 | 40 | 39 | 39 | 38 | 37 | 36 | 37 |
| Occupational Therapist in Private Practice | 60 | 54 | 51 | 51 | 49 | 48 | 46 | 45 | 46 |
| Ophthalmology | 49 | 48 | 48 | 47 | 46 | 46 | 46 | 45 | 44 |
| Optometry | 76 | 75 | 73 | 71 | 69 | 67 | 66 | 64 | 63 |
| Oral Surgery (Dentist only) | 46 | 45 | 43 | 42 | 42 | 43 | 44 | 45 | 45 |
| Orthopedic Surgery | 30 | 29 | 28 | 28 | 27 | 27 | 26 | 26 | 27 |
| Osteopathic Manipulative Medicine | 50 | 46 | 44 | 43 | 41 | 41 | 40 | 39 | 40 |
| Otolaryngology | 42 | 41 | 39 | 38 | 37 | 36 | 36 | 35 | 35 |
| Pain Management | 30 | 29 | 27 | 26 | 24 | 24 | 24 | 23 | 24 |
| Pathology | 31 | 29 | 28 | 28 | 29 | 29 | 29 | 28 | 28 |
| Pediatric Medicine | 44 | 41 | 42 | 40 | 39 | 39 | 40 | 38 | 37 |
| Peripheral Vascular Disease | 33 | 32 | 27 | 26 | 29 | 31 | 33 | 35 | 34 |
| Physical Medicine and Rehabilitation | 44 | 42 | 40 | 39 | 37 | 36 | 35 | 34 | 34 |
| Physical Therapist in Private Practice | 57 | 52 | 50 | 49 | 48 | 47 | 46 | 44 | 44 |
| Physician Assistant | 26 | 25 | 25 | 25 | 25 | 25 | 25 | 25 | 25 |
| Plastic and Reconstructive Surgery | 31 | 30 | 29 | 28 | 27 | 26 | 26 | 25 | 24 |
| Podiatry | 61 | 61 | 59 | 57 | 55 | 54 | 53 | 53 | 53 |
| Preventive Medicine | 42 | 39 | 38 | 38 | 35 | 33 | 36 | 36 | 37 |
| Psychiatry | 54 | 55 | 56 | 55 | 53 | 51 | 51 | 49 | 49 |
| Psychologist, Clinical | 53 | 57 | 65 | 64 | 59 | 58 | 57 | 56 | 56 |
| Pulmonary Disease | 49 | 48 | 47 | 46 | 44 | 43 | 43 | 42 | 41 |
| Radiation Oncology | 29 | 27 | 28 | 27 | 26 | 26 | 26 | 26 | 26 |
| Registered Dietitian or Nutrition Profess | 54 | 53 | 52 | 49 | 48 | 47 | 45 | 46 | 45 |
| Rheumatology | 54 | 55 | 54 | 55 | 55 | 55 | 54 | 50 | 49 |
| Sleep Medicine | 29 | 36 | 37 | 37 | 37 | 36 | 37 | 35 | 36 |
| Speech Language Pathologist | 63 | 62 | 60 | 58 | 57 | 52 | 52 | 53 | 52 |
| Sports Medicine | 32 | 33 | 31 | 32 | 32 | 32 | 31 | 33 | 32 |
| Surgical Oncology | 29 | 29 | 28 | 28 | 27 | 27 | 27 | 26 | 26 |
| Thoracic Surgery | 27 | 26 | 27 | 27 | 27 | 27 | 27 | 26 | 26 |
| Urology | 37 | 36 | 36 | 36 | 35 | 34 | 34 | 33 | 33 |
| Vascular Surgery | 30 | 29 | 29 | 29 | 29 | 30 | 30 | 30 | 29 |
| TOTAL | 39 | 38 | 37 | 37 | 36 | 35 | 34 | 33 | 33 |

Source of data: CMS public access data tabulated by Denniston Data Inc. at PRS [16-17]

**Table 4:** Averaged Provider Data For Year 2018 In Terms Of Billed & Approved Dollars, Codes & Services Billed, And Patients Cared.

| Specialty Name | Number of Providers | Billed \$ Per Provider | Approved \$ Per Provider | Codes Per Provider | Services Per Provider | Patients Per Provider |
| --- | --- | --- | --- | --- | --- | --- |
| Addiction Medicine | 199 | 124,115 | 59,971 | 14 | 915 | 117 |
| Allergy/ Immunology | 3,266 | 233,125 | 121,353 | 24 | 6,116 | 192 |
| Anesthesiology | 34,662 | 451,820 | 61,417 | 53 | 568 | 257 |
| Anesthesiology Assistant | 1,646 | 225,925 | 24,696 | 51 | 186 | 178 |
| Audiologist | 6,631 | 30,927 | 10,710 | 7 | 321 | 151 |
| Cardiac Surgery | 1,264 | 865,009 | 228,907 | 58 | 852 | 230 |
| Cardiology | 19,743 | 789,383 | 283,924 | 48 | 3,670 | 1,080 |
| Certified Clinical Nurse Specialist | 1,914 | 84,248 | 36,525 | 10 | 643 | 138 |
| Certified Nurse Midwife | 500 | 17,507 | 8,029 | 19 | 174 | 64 |
| Certified Registered Nurse Anesthetist | 36,836 | 249,727 | 33,096 | 42 | 225 | 202 |
| Chiropractic | 34,460 | 29,583 | 20,997 | 2 | 571 | 63 |
| Clinical Cardiac Electrophysiology | 2,119 | 923,284 | 305,847 | 73 | 3,803 | 1,175 |
| Colorectal Surgery (Proctology) | 1,458 | 383,334 | 113,415 | 63 | 595 | 223 |
| Critical Care (Intensivists) | 3,434 | 310,868 | 106,731 | 25 | 888 | 265 |
| Dermatology | 11,658 | 659,180 | 314,397 | 52 | 4,437 | 729 |
| Diagnostic Radiology | 29,941 | 732,038 | 179,512 | 119 | 6,328 | 2,132 |
| Emergency Medicine | 43,581 | 408,311 | 72,047 | 24 | 680 | 407 |
| Endocrinology | 5,693 | 237,872 | 109,662 | 22 | 3,060 | 364 |
| Family Practice | 81,452 | 177,183 | 82,502 | 41 | 1,941 | 258 |
| Gastroenterology | 13,444 | 522,201 | 141,843 | 40 | 1,822 | 434 |
| General Practice | 4,683 | 165,082 | 81,526 | 27 | 1,561 | 192 |
| General Surgery | 19,039 | 356,469 | 104,691 | 61 | 665 | 210 |
| Geriatric Medicine | 1,814 | 230,007 | 110,592 | 25 | 1,404 | 297 |
| Geriatric Psychiatry | 198 | 195,214 | 100,400 | 11 | 1,127 | 257 |
| Gynecological Oncology | 940 | 517,434 | 157,244 | 54 | 5,338 | 178 |
| Hand Surgery | 1,463 | 587,935 | 165,404 | 84 | 1,953 | 361 |
| Hematology | 745 | 954,120 | 363,258 | 29 | 19,615 | 239 |
| Hematology-Oncology | 8,193 | 2,302,437 | 923,016 | 56 | 53,072 | 383 |
| Hospice and Palliative Care | 1,190 | 111,878 | 47,139 | 14 | 592 | 159 |
| Infectious Disease | 5,699 | 313,075 | 139,391 | 18 | 9,072 | 291 |
| Internal Medicine | 90,516 | 271,503 | 121,616 | 30 | 2,368 | 327 |
| Interventional Pain Management | 1,633 | 1,251,080 | 307,658 | 48 | 5,637 | 420 |
| Interventional Radiology | 1,729 | 1,115,785 | 257,624 | 126 | 3,988 | 974 |
| Licensed Clinical Social Worker | 18,719 | 45,099 | 24,369 | 4 | 338 | 35 |
| Maxillofacial Surgery | 673 | 81,615 | 24,905 | 27 | 172 | 71 |
| Medical Oncology | 3,237 | 1,856,624 | 714,078 | 48 | 38,224 | 308 |
| Nephrology | 8,606 | 598,446 | 266,147 | 26 | 3,633 | 459 |
| Neurology | 14,138 | 377,582 | 143,793 | 24 | 3,635 | 346 |
| Neuropsychiatry | 117 | 229,930 | 85,762 | 17 | 1,648 | 217 |
| Neurosurgery | 4,563 | 858,540 | 176,111 | 52 | 675 | 223 |
| Nuclear Medicine | 532 | 730,381 | 241,032 | 45 | 2,386 | 940 |
| Nurse Practitioner | 116,881 | 100,875 | 36,976 | 20 | 824 | 176 |
| Obstetrics & Gynecology | 21,548 | 71,865 | 26,733 | 27 | 473 | 111 |
| Occupational Therapist in Private Practice | 5,718 | 101,326 | 47,102 | 10 | 1,599 | 54 |
| Ophthalmology | 17,421 | 1,186,064 | 545,498 | 34 | 3,517 | 730 |
| Optometry | 28,611 | 68,727 | 45,345 | 14 | 770 | 249 |
| Oral Surgery (Dentist only) | 1,102 | 120,296 | 53,357 | 17 | 251 | 83 |
| Orthopedic Surgery | 20,892 | 729,836 | 191,900 | 71 | 2,549 | 349 |
| Osteopathic Manipulative Medicine | 614 | 221,225 | 87,507 | 25 | 2,033 | 182 |
| Otolaryngology | 8,645 | 400,238 | 144,480 | 54 | 1,857 | 412 |
| Pain Management | 2,252 | 953,093 | 229,145 | 45 | 3,790 | 365 |
| Pathology | 11,749 | 405,308 | 115,774 | 23 | 2,666 | 837 |
| Pediatric Medicine | 1,533 | 127,543 | 50,983 | 25 | 1,618 | 155 |
| Peripheral Vascular Disease | 53 | 1,273,339 | 425,902 | 55 | 2,105 | 770 |
| Physical Medicine and Rehabilitation | 7,844 | 459,162 | 159,969 | 27 | 3,341 | 314 |
| Physical Therapist in Private Practice | 54,160 | 137,626 | 62,834 | 10 | 2,272 | 81 |
| Physician Assistant | 70,272 | 142,330 | 35,170 | 27 | 773 | 195 |
| Plastic and Reconstructive Surgery | 3,671 | 405,975 | 104,750 | 65 | 630 | 142 |
| Podiatry | 15,155 | 264,216 | 140,485 | 41 | 2,253 | 459 |
| Preventive Medicine | 289 | 175,051 | 62,273 | 22 | 2,136 | 165 |
| Psychiatry | 21,008 | 105,322 | 53,330 | 9 | 670 | 128 |
| Psychologist, Clinical | 15,022 | 80,316 | 45,483 | 5 | 488 | 61 |
| Pulmonary Disease | 9,971 | 417,488 | 178,227 | 33 | 2,146 | 500 |
| Radiation Oncology | 4,548 | 1,655,286 | 432,201 | 35 | 3,621 | 243 |
| Registered Dietitian or Nutrition Profess | 1,871 | 19,861 | 8,994 | 3 | 305 | 59 |
| Rheumatology | 4,607 | 1,169,727 | 631,123 | 36 | 26,570 | 359 |
| Sleep Medicine | 466 | 312,166 | 115,592 | 15 | 1,012 | 396 |
| Speech Language Pathologist | 975 | 86,085 | 44,785 | 7 | 686 | 50 |
| Sports Medicine | 1,193 | 353,500 | 111,113 | 43 | 2,785 | 277 |
| Surgical Oncology | 919 | 374,828 | 102,058 | 49 | 665 | 175 |
| Thoracic Surgery | 2,128 | 626,502 | 168,251 | 56 | 535 | 181 |
| Urology | 8,838 | 761,236 | 256,435 | 73 | 8,370 | 591 |
| Vascular Surgery | 3,248 | 1,251,151 | 374,639 | 93 | 2,192 | 585 |
| TOTAL | 985,532 | 326,690 | 112,080 | 33 | 2,467 | 338 |

Source of data: CMS public access data tabulated by Denniston Data Inc. at PRS [16-17]

**Table 5:** Averaged Provider Data For Year 2019 In Terms Of Billed & Approved Dollars, Codes & Services Billed, And Patients Cared.

| Specialty Name | Number of Providers | Billed \$ Per Provider | Approved \$ Per Provider | Codes Per Provider | Services Per Provider | Patients Per Provider |
| --- | --- | --- | --- | --- | --- | --- |
| Addiction Medicine | 214 | 117,922 | 54,133 | 13 | 761 | 101 |
| Allergy/ Immunology | 3,266 | 265,780 | 131,416 | 24 | 6,506 | 195 |
| Anesthesiology | 35,004 | 463,278 | 59,720 | 53 | 556 | 254 |
| Anesthesiology Assistant | 1,777 | 226,878 | 23,979 | 51 | 184 | 177 |
| Audiologist | 6,852 | 31,328 | 10,721 | 7 | 321 | 152 |
| Cardiac Surgery | 1,220 | 866,534 | 223,972 | 56 | 742 | 220 |
| Cardiology | 19,654 | 799,803 | 282,818 | 47 | 3,614 | 1,081 |
| Certified Clinical Nurse Specialist | 1,857 | 84,666 | 35,387 | 10 | 615 | 137 |
| Certified Nurse Midwife | 525 | 17,921 | 7,871 | 19 | 178 | 62 |
| Certified Registered Nurse Anesthetist | 38,190 | 258,265 | 32,492 | 4 | 224 | 200 |
| Chiropractic | 34,309 | 30,432 | 21,522 | 2 | 575 | 64 |
| Clinical Cardiac Electrophysiology | 2,207 | 1,026,240 | 320,852 | 72 | 4,000 | 1,215 |
| Colorectal Surgery (Proctology) | 1,473 | 383,639 | 112,476 | 62 | 596 | 222 |
| Critical Care (Intensivists) | 3,712 | 313,141 | 102,112 | 25 | 842 | 262 |
| Dermatology | 11,899 | 692,544 | 319,609 | 54 | 4,533 | 730 |
| Diagnostic Radiology | 30,262 | 768,872 | 181,825 | 117 | 6,685 | 2,136 |
| Emergency Medicine | 44,458 | 414,484 | 68,975 | 24 | 652 | 393 |
| Endocrinology | 5,825 | 244,464 | 107,915 | 21 | 2,982 | 361 |
| Family Practice | 81,425 | 181,423 | 80,717 | 40 | 1,884 | 254 |
| Gastroenterology | 13,619 | 536,517 | 141,583 | 40 | 1,931 | 430 |
| General Practice | 4,490 | 165,018 | 79,663 | 27 | 1,561 | 188 |
| General Surgery | 19,156 | 355,987 | 102,647 | 59 | 646 | 207 |
| Geriatric Medicine | 1,847 | 233,979 | 107,000 | 25 | 1,444 | 285 |
| Geriatric Psychiatry | 192 | 193,117 | 96,592 | 11 | 1,087 | 239 |
| Gynecological Oncology | 947 | 545,821 | 163,592 | 53 | 5,730 | 176 |
| Hand Surgery | 1,516 | 615,457 | 171,208 | 84 | 2,055 | 377 |
| Hematology | 770 | 983,195 | 348,751 | 28 | 19,909 | 238 |
| Hematology-Oncology | 8,287 | 2,455,306 | 944,111 | 56 | 54,558 | 385 |
| Hospice and Palliative Care | 1,308 | 116,177 | 46,951 | 14 | 557 | 157 |
| Infectious Disease | 5,815 | 316,151 | 136,069 | 18 | 9,316 | 282 |
| Internal Medicine | 89,808 | 277,294 | 118,476 | 30 | 2,368 | 320 |
| Interventional Pain Management | 1,611 | 1,263,581 | 305,869 | 48 | 5,738 | 410 |
| Interventional Radiology | 1,906 | 1,149,558 | 266,756 | 126 | 4,023 | 977 |
| Licensed Clinical Social Worker | 19,373 | 46,399 | 24,645 | 4 | 334 | 35 |
| Maxillofacial Surgery | 691 | 82,883 | 25,102 | 26 | 182 | 69 |
| Medical Oncology | 3,315 | 2,014,515 | 744,880 | 48 | 39,807 | 311 |
| Nephrology | 8,783 | 594,165 | 258,062 | 25 | 3,465 | 456 |
| Neurology | 14,451 | 391,889 | 142,408 | 23 | 3,709 | 339 |
| Neuropsychiatry | 122 | 233,654 | 80,999 | 17 | 1,556 | 220 |
| Neurosurgery | 4,640 | 845,452 | 173,262 | 51 | 659 | 220 |
| Nuclear Medicine | 535 | 765,287 | 242,921 | 46 | 2,648 | 974 |
| Nurse Practitioner | 130,561 | 107,240 | 37,910 | 20 | 894 | 175 |
| Obstetrics & Gynecology | 21,189 | 74,316 | 26,989 | 27 | 484 | 110 |
| Occupational Therapist in Private Practice | 6,431 | 107,351 | 48,191 | 10 | 1,657 | 55 |
| Ophthalmology | 17,494 | 1,247,569 | 558,946 | 34 | 3,545 | 721 |
| Optometry | 29,216 | 71,286 | 45,936 | 14 | 786 | 251 |
| Oral Surgery (Dentist only) | 1,077 | 117,481 | 53,404 | 17 | 250 | 79 |
| Orthopedic Surgery | 20,918 | 741,691 | 195,874 | 69 | 2,611 | 348 |
| Osteopathic Manipulative Medicine | 630 | 228,506 | 89,638 | 24 | 2,106 | 177 |
| Otolaryngology | 8,696 | 411,282 | 144,400 | 53 | 1,863 | 410 |
| Pain Management | 2,403 | 984,972 | 229,887 | 45 | 3,971 | 363 |
| Pathology | 11,735 | 422,664 | 116,753 | 23 | 2,796 | 867 |
| Pediatric Medicine | 1,539 | 139,037 | 52,708 | 25 | 1,589 | 160 |
| Peripheral Vascular Disease | 53 | 1,311,090 | 454,307 | 57 | 2,108 | 801 |
| Physical Medicine and Rehabilitation | 7,905 | 477,362 | 160,470 | 27 | 3,494 | 314 |
| Physical Therapist in Private Practice | 58,351 | 144,214 | 63,703 | 10 | 2,327 | 82 |
| Physician Assistant | 75,704 | 145,706 | 35,877 | 27 | 820 | 196 |
| Plastic and Reconstructive Surgery | 3,690 | 419,663 | 104,181 | 64 | 634 | 141 |
| Podiatry | 15,212 | 269,380 | 141,757 | 41 | 2,218 | 455 |
| Preventive Medicine | 263 | 177,940 | 64,127 | 22 | 2,560 | 164 |
| Psychiatry | 20,822 | 105,494 | 51,916 | 8 | 646 | 121 |
| Psychologist, Clinical | 14,799 | 82,612 | 45,930 | 6 | 508 | 61 |
| Pulmonary Disease | 10,068 | 413,936 | 171,876 | 33 | 2,097 | 490 |
| Radiation Oncology | 4,612 | 1,693,500 | 432,901 | 35 | 3,661 | 245 |
| Registered Dietitian or Nutrition Profess | 1,933 | 21,409 | 9,766 | 3 | 316 | 59 |
| Rheumatology | 4,699 | 1,309,659 | 651,147 | 36 | 29,611 | 357 |
| Sleep Medicine | 535 | 302,811 | 107,067 | 15 | 932 | 387 |
| Speech Language Pathologist | 1,145 | 90,637 | 47,853 | 7 | 734 | 52 |
| Sports Medicine | 1,295 | 371,214 | 121,090 | 43 | 2,880 | 286 |
| Surgical Oncology | 966 | 385,721 | 99,750 | 49 | 665 | 174 |
| Thoracic Surgery | 2,166 | 624,081 | 163,611 | 54 | 495 | 175 |
| Urology | 8,891 | 785,746 | 255,538 | 72 | 8,373 | 587 |
| Vascular Surgery | 3,364 | 1,303,114 | 384,816 | 91 | 2,219 | 579 |
| TOTAL | 1,015,673 | 333,849 | 110,876 | 31 | 2,500 | 333 |

Source of data: CMS public access data tabulated by Denniston Data Inc. at PRS [16-17]

**Table 6:** Averaged Provider Data For Year 2020 In Terms Of Billed & Approved Dollars, Codes & Services Billed, And Patients Cared.

| Specialty Name | Number of Providers | Billed \$ Per Provider | Approved \$ Per Provider | Codes Per Provider | Services Per Provider | Patients Per Provider |
| --- | --- | --- | --- | --- | --- | --- |
| Addiction Medicine | 201 | 97,381 | 44,747 | 14 | 573 | 94 |
| Allergy/ Immunology | 3,181 | 279,042 | 135,521 | 24 | 6,215 | 181 |
| Anesthesiology | 33,471 | 418,616 | 53,657 | 51 | 490 | 219 |
| Anesthesiology Assistant | 1,683 | 216,572 | 22,783 | 53 | 173 | 166 |
| Audiologist | 6,684 | 26,554 | 8,693 | 6 | 255 | 121 |
| Cardiac Surgery | 1,126 | 750,079 | 187,164 | 53 | 544 | 184 |
| Cardiology | 19,419 | 696,773 | 246,195 | 47 | 3,091 | 937 |
| Certified Clinical Nurse Specialist | 1,679 | 83,552 | 34,297 | 12 | 597 | 127 |
| Certified Nurse Midwife | 411 | 17,926 | 8,337 | 21 | 169 | 62 |
| Certified Registered Nurse Anesthetist | 35,666 | 243,818 | 29,886 | 42 | 202 | 179 |
| Chiropractic | 32,395 | 27,164 | 19,205 | 2 | 505 | 58 |
| Clinical Cardiac Electrophysiology | 2,293 | 941,206 | 294,013 | 71 | 3,636 | 1,092 |
| Colorectal Surgery (Proctology) | 1,460 | 326,883 | 97,647 | 60 | 509 | 187 |
| Critical Care (Intensivists) | 3,978 | 295,435 | 95,063 | 24 | 729 | 225 |
| Dermatology | 11,930 | 625,715 | 284,237 | 52 | 3,861 | 637 |
| Diagnostic Radiology | 30,362 | 700,133 | 160,891 | 110 | 6,010 | 1,794 |
| Emergency Medicine | 44,583 | 346,219 | 57,592 | 22 | 524 | 316 |
| Endocrinology | 5,836 | 224,541 | 98,634 | 23 | 2,700 | 333 |
| Family Practice | 79,480 | 165,029 | 73,021 | 39 | 1,661 | 234 |
| Gastroenterology | 13,612 | 459,387 | 123,511 | 40 | 1,827 | 364 |
| General Practice | 3,965 | 163,549 | 77,786 | 27 | 1,455 | 176 |
| General Surgery | 18,812 | 315,071 | 90,328 | 55 | 564 | 177 |
| Geriatric Medicine | 1,829 | 208,941 | 95,721 | 26 | 1,288 | 249 |
| Geriatric Psychiatry | 179 | 182,284 | 89,689 | 12 | 1,003 | 206 |
| Gynecological Oncology | 943 | 552,674 | 163,344 | 52 | 5,958 | 158 |
| Hand Surgery | 1,510 | 544,049 | 151,774 | 80 | 1,785 | 332 |
| Hematology | 778 | 920,887 | 330,699 | 28 | 18,484 | 215 |
| Hematology-Oncology | 8,354 | 2,478,814 | 942,171 | 56 | 51,509 | 354 |
| Hospice and Palliative Care | 1,394 | 112,595 | 45,918 | 15 | 502 | 145 |
| Infectious Disease | 5,844 | 320,210 | 137,338 | 18 | 9,500 | 275 |
| Internal Medicine | 87,738 | 256,023 | 107,730 | 29 | 2,104 | 288 |
| Interventional Pain Management | 1,566 | 1,155,880 | 281,568 | 49 | 5,222 | 366 |
| Interventional Radiology | 2,002 | 1,085,597 | 250,552 | 122 | 3,718 | 855 |
| Licensed Clinical Social Worker | 17,052 | 48,634 | 25,661 | 5 | 333 | 33 |
| Maxillofacial Surgery | 528 | 94,087 | 28,319 | 29 | 187 | 68 |
| Medical Oncology | 3,433 | 1,984,677 | 719,613 | 48 | 36,465 | 282 |
| Nephrology | 8,902 | 563,703 | 243,066 | 26 | 2,974 | 420 |
| Neurology | 14,555 | 346,450 | 124,233 | 24 | 3,370 | 292 |
| Neuropsychiatry | 118 | 179,683 | 75,010 | 19 | 1,588 | 202 |
| Neurosurgery | 4,626 | 764,072 | 153,794 | 51 | 580 | 190 |
| Nuclear Medicine | 517 | 805,348 | 252,556 | 43 | 3,071 | 852 |
| Nurse Practitioner | 137,536 | 104,926 | 37,594 | 20 | 859 | 159 |
| Obstetrics & Gynecology | 19,060 | 69,237 | 25,396 | 27 | 461 | 101 |
| Occupational Therapist in Private Practice | 6,667 | 94,735 | 43,426 | 9 | 1,460 | 47 |
| Ophthalmology | 17,192 | 1,127,572 | 501,695 | 33 | 2,974 | 614 |
| Optometry | 28,228 | 60,292 | 38,078 | 14 | 627 | 214 |
| Oral Surgery (Dentist only) | 879 | 128,721 | 58,183 | 19 | 252 | 74 |
| Orthopedic Surgery | 20,695 | 645,354 | 171,201 | 66 | 2,302 | 297 |
| Osteopathic Manipulative Medicine | 609 | 210,332 | 84,812 | 24 | 1,908 | 164 |
| Otolaryngology | 8,606 | 337,627 | 119,307 | 50 | 1,532 | 341 |
| Pain Management | 2,461 | 865,138 | 209,631 | 46 | 3,589 | 324 |
| Pathology | 11,686 | 377,874 | 104,974 | 22 | 2,496 | 747 |
| Pediatric Medicine | 1,532 | 122,514 | 45,268 | 23 | 1,865 | 148 |
| Peripheral Vascular Disease | 52 | 1,156,069 | 389,651 | 57 | 1,807 | 757 |
| Physical Medicine and Rehabilitation | 7,865 | 424,767 | 142,506 | 27 | 3,091 | 270 |
| Physical Therapist in Private Practice | 58,835 | 117,646 | 51,775 | 10 | 1,856 | 64 |
| Physician Assistant | 77,071 | 132,685 | 33,207 | 26 | 752 | 174 |
| Plastic and Reconstructive Surgery | 3,347 | 419,669 | 102,784 | 63 | 595 | 128 |
| Podiatry | 15,149 | 247,945 | 131,164 | 40 | 1,842 | 393 |
| Preventive Medicine | 238 | 169,234 | 62,538 | 22 | 1,534 | 154 |
| Psychiatry | 19,761 | 99,773 | 48,611 | 9 | 600 | 107 |
| Psychologist, Clinical | 13,106 | 83,698 | 47,182 | 6 | 493 | 57 |
| Pulmonary Disease | 10,066 | 375,072 | 153,604 | 33 | 1,789 | 422 |
| Radiation Oncology | 4,646 | 1,554,748 | 400,880 | 35 | 3,298 | 216 |
| Registered Dietitian or Nutrition Profess | 1,758 | 19,754 | 8,955 | 4 | 275 | 55 |
| Rheumatology | 4,722 | 1,368,590 | 667,160 | 37 | 32,044 | 326 |
| Sleep Medicine | 620 | 226,957 | 81,992 | 17 | 774 | 341 |
| Speech Language Pathologist | 1,126 | 80,981 | 42,224 | 7 | 698 | 45 |
| Sports Medicine | 1,360 | 331,683 | 105,291 | 42 | 2,631 | 245 |
| Surgical Oncology | 964 | 349,936 | 90,226 | 47 | 506 | 152 |
| Thoracic Surgery | 2,140 | 560,850 | 146,425 | 53 | 431 | 153 |
| Urology | 8,849 | 707,240 | 230,277 | 70 | 7,721 | 521 |
| Vascular Surgery | 3,380 | 1,177,422 | 345,191 | 89 | 1,987 | 496 |
| TOTAL | 1,004,301 | 306,326 | 101,673 | 32 | 2,305 | 291 |

Source of data: CMS public access data tabulated by Denniston Data Inc. at PRS [16-17]

**Table 7:** Metaphorical Comparison Among Specialties Deciphering CMS Contributions To Annual Mean Wages Of Providers.

| Specialty Name | Annual Approved \$<br>Per PRS (CMS)<br>Averaged 2018-2020 | Annual Providers n<br>Per PRS (CMS)<br>Averaged 2018-2020 | Annual \$ Per Provider<br>Per PRS (CMS)<br>Averaged 2018-2020 | Total Employment<br>(Number of Providers)<br>Per May 2021 BLS | Provider's Annual<br>Mean Wage<br>Per May 2021 BLS | Provider's Wage Per<br>BLS (-) Provider's \$ Per<br>PRS (CMS) |
| --- | --- | --- | --- | --- | --- | --- |
| Anesthesiology | 2,005,076,417 | 34,379 | 58,323 | 31,130 | 331,190 | 272,867 |
| Audiologist | 67,528,772 | 6,722 | 10,045 | 13,240 | 86,050 | 76,005 |
| Cardiology | 5,314,965,425 | 19,605 | 271,098 | 18,610 | 353,970 | 82,872 |
| Certified Nurse Midwife | 3,857,819 | 479 | 8,060 | 7,750 | 114,210 | 106,150 |
| Certified Registered Nurse Anesthetist | 1,175,293,689 | 36,897 | 31,853 | 43,950 | 202,470 | 170,617 |
| Chiropractic | 694,705,400 | 33,721 | 20,601 | 35,810 | 81,240 | 60,639 |
| Dermatology | 3,619,737,430 | 11,829 | 306,005 | 9,230 | 302,740 | -3,265 |
| Emergency Medicine | 2,924,671,354 | 44,207 | 66,158 | 36,180 | 310,640 | 244,482 |
| Family Practice | 6,365,367,172 | 80,786 | 78,793 | 102,930 | 235,930 | 157,137 |
| Internal Medicine | 10,366,800,695 | 89,354 | 116,019 | 58,260 | 242,190 | 126,171 |
| Neurology | 1,966,365,134 | 14,381 | 136,730 | 7,120 | 267,660 | 130,930 |
| Nurse Practitioner | 4,813,934,181 | 128,326 | 37,513 | 234,690 | 118,040 | 80,527 |
| Obstetrics & Gynecology | 543,991,745 | 20,599 | 26,409 | 21,570 | 296,210 | 269,801 |
| Occupational Therapist in Private Practice | 289,590,953 | 6,272 | 46,172 | 127,830 | 89,470 | 43,298 |
| Ophthalmology | 9,302,154,273 | 17,369 | 535,561 | 11,610 | 270,090 | -265,471 |
| Optometry | 1,238,093,815 | 28,685 | 43,162 | 38,720 | 125,440 | 82,278 |
| Orthopedic Surgery | 3,883,155,760 | 20,835 | 186,377 | 16,260 | 306,220 | 119,843 |
| Pathology | 1,319,017,670 | 11,723 | 112,512 | 11,010 | 267,180 | 154,668 |
| Pediatric Medicine | 76,208,182 | 1,535 | 49,658 | 33,620 | 198,420 | 148,762 |
| Physical Therapist in Private Practice | 3,388,803,924 | 57,115 | 59,333 | 225,350 | 92,920 | 33,587 |
| Physician Assistant | 2,582,262,028 | 74,349 | 34,732 | 132,940 | 119,460 | 84,728 |
| Podiatry | 2,090,822,402 | 15,172 | 137,808 | 8,840 | 158,380 | 20,572 |
| Psychiatry | 1,053,980,702 | 20,530 | 51,338 | 25,520 | 249,760 | 198,422 |

Sources of data: CMS public access data tabulated by Denniston Data Inc. at PRS [16-17]; and public access BLS OEWS May 2021 Estimates [10-11]

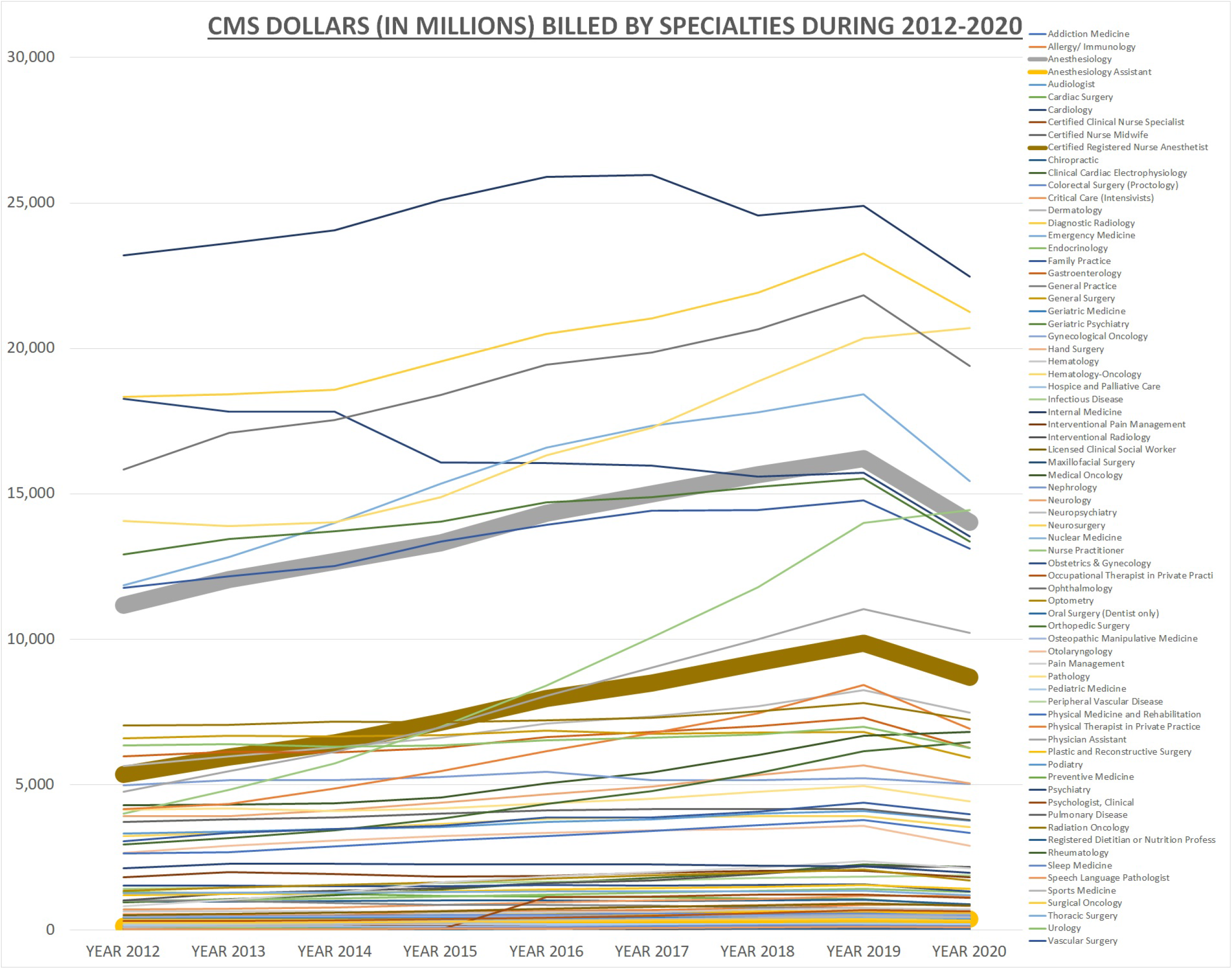

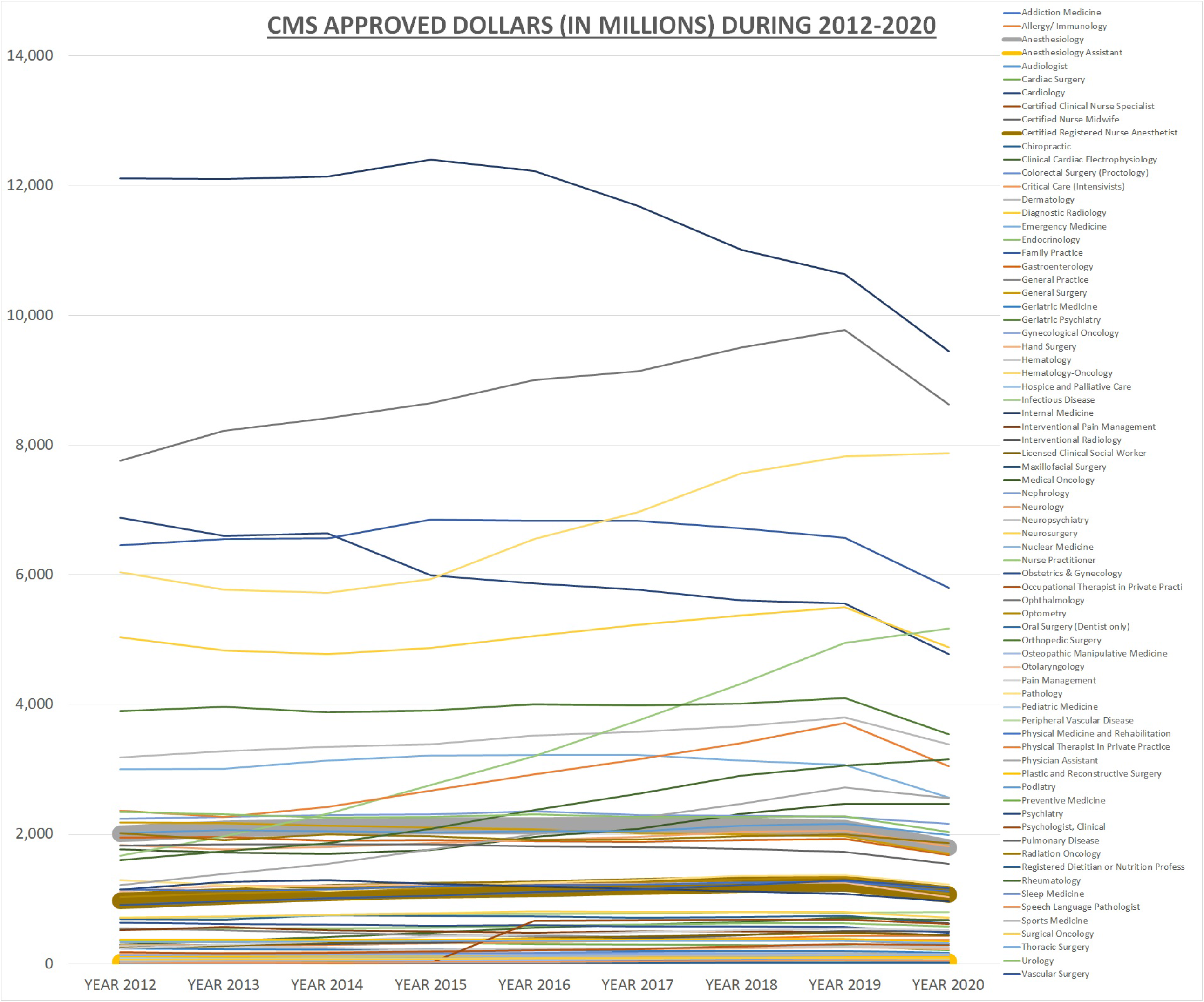

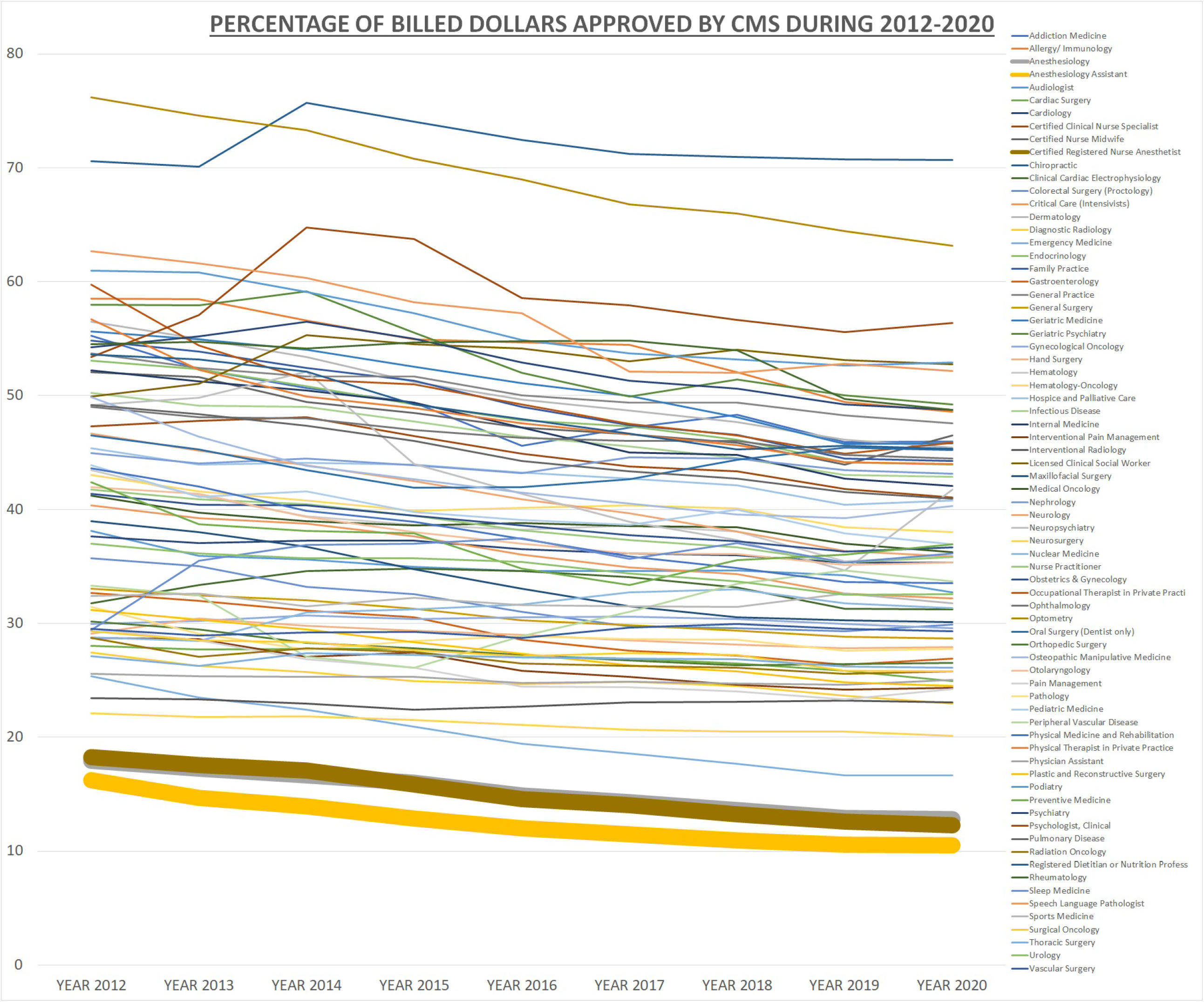

## Discussion

In 2020 [12], Medicare and Medicaid collectively were the source of funds for 36.4% of 4.1 trillion dollars as NHE making CMS the largest healthcare payer and thus its approved dollars representing national scenario in terms of reimbursements for healthcare providers. Although private health insurance has been the next major source of funds for NHE, private health insurance has been known to approve dollars as higher percentages of billed dollars as compared to CMS [19], and thus maybe covering far smaller number of populations in terms of codes and services billed for patients cared than CMS despite being the source of funds for 27.9% NHE in 2020. Herein the presented tables and figures clearly show that anesthesia providers’ billed dollars are getting approved at less than 20% with most of the times these approved percentages hovering around low 10s. Thus, it may appear that anesthesiologists, who are nationally ranked second to only cardiologists in terms of annual mean wages per BLS OEWS May 2021 estimates [5], may be drawing their annual wages mostly from non-CMS payers and direct payments from healthcare institutions like hospitals [20].

Herein the following questions arise:

- Are anesthesia providers being undervalued by CMS because they mostly work as teams wherein anesthesiologists are submitting bills concurrently with either CRNAs and/or anesthesiology assistants [21-22]?
- Are the billed dollars submitted by anesthesiologists personally performing anesthesia services higher than the billed dollars submitted by anesthesiologists medically directing anesthesia services [23]?
- Are the billed dollars submitted by medically directed CRNAs during anesthesia services lower than the billed dollars submitted by CRNAs performing anesthesia services without medical direction [23]?

Besides the discrepancy when compared to other online data [24], the discrepancy in annual providers’ number per PRS versus total employment (number of providers) per BLS may be because of BLS OEWS survey excluding self-employed, owners and partners in unincorporated firms which may be a lot of healthcare providers especially physicians [4], while PRS data excluding those providers who had neither submitted billed dollars to CMS nor received approved dollars from CMS. Thereafter, difference between provider’s wage per BLS versus provider’s approved dollars per PRS becoming negative in dermatology and ophthalmology may be indicating that BLS estimates may have excluded too many providers from these specialties due to potential preponderance of dermatologists and ophthalmologists being self-employed, owners and partners in unincorporated firms. Even in other specialties too, the difference between provider’s wage per BLS versus provider’s approved dollars per PRS may be just indicating, without actually quantifying, the contributions by non-CMS approved dollars to providers practicing in those specialties considering that most, if not all healthcare providers, do not directly receive annually approved dollars (CMS as well as non-CMS) as their annual wage because of their practice groups and their employers along with their administrative costs in the middle which have to keep some for their own survival while collecting on behalf of healthcare providers not only the directly approved dollars from healthcare payers like CMS as well as non-CMS but also the direct payments from healthcare institutions like hospitals when the directly approved dollars from healthcare payers are well-below healthcare providers’ expectations for their annual wage.

## Conclusion

The bottom line is that unlike maybe all other professionals including lawyers whose bills may be expected to be paid in full, healthcare providers especially anesthesia providers are being paid a very small fraction of their billed dollars and yet anesthesiologists averaging almost top 1% wages (99th percentile [1]) nationally with CRNAs averaging top 5% wages (95th percentile [1]) nationally raise the question whether healthcare providers are overvaluing themselves when billing healthcare payers who are undervaluing healthcare providers when approving only percentages of billed dollars thus driving healthcare providers to further overvalue themselves for drawing fair wages well-above minimum wages and maybe even drawing living wages for themselves in this endlessly dynamic tug-of-war to balance valuation and pricing [25-26]. Moreover, considering that Cost-to-Charge Ratios for hospital care as well as ratios of approved dollars to billed dollars (percentage of billed dollars approved) for healthcare providers may be extrapolated to be hovering around one-thirds overall nationally [27], it can only be wondered whether NHE would have tripled in 2020 to >12 trillion dollars and thus share of NHE in GDP would have more than doubled in 2020 to >41.8% instead of share of NHE in GDP tripling to almost 60% because GDP in 2020 would have ironically grown from 20.8 trillion dollars to >28.7 trillion dollars on the backs of stupendous growth in healthcare dollars if hospital care charges and healthcare provider bills were designed to be 100% approved by healthcare payers.

## Supporting information

IRB

## Data Availability

All data produced in the present work are contained in the manuscript.
Moreover, data produced in the present work can be sourced to raw public access datasets available from Data Availability Links.

https://www.bls.gov/oes/current/oes_nat.htm

https://www.providerrankings.com/

https://www.bls.gov/oes/special.requests/oesm21nat.zip

https://dennistondata.com/source-of-data/

https://data.cms.gov/provider-summary-by-type-of-service/medicare-physician-other-practitioners

## Acknowledgement

The author is indebted to Thomas Farrell Denniston, Account Executive, Denniston Data Inc., Corpus Christi, Texas, United States, for providing cumulative deidentified data tables for years 2012-2020 used in this presented project.

